# Parent co-Designed Drug Information for parents and Guardians Taking Neonates home (PADDINGToN) a mixed method approach to defining and evaluating information resources

**DOI:** 10.1101/2022.07.10.22277472

**Authors:** Andrea Gill, Louise Bracken, Catrin Barker, Neil Caldwell, Brian Cleary, Naomi McCallion, Stephen Morris, Elaine Neary, Mark Turner, Matthew Peak, Fiona O’Brien

## Abstract

**Background:** A study of premature babies, less than 32 weeks gestation, found that the median number of medicines per patient at discharge was 2.5 (range 2 to 7), with 28% of babies discharged on more than 3 medicines (1). Medication administration to infants can cause anxiety and concern for parents/carers who worry about making mistakes. A systematic review of carers for all ages has estimated the administration error rate at home to be between 2 and 33% (2) while a rate of 66.3% was recorded in medication administered to premature infants where parents were responsible for administration (3). It has also been reported that up to 90% of carers will administer a medicine incorrectly at some point (2).

The aim of this study is to work with healthcare professionals (HCPs) and parents/carers to co-design resources aimed at improving medication safety and reducing parental anxiety for those giving medications to neonates at home.

**Methods:** *Work Package 1: Project management and co-ordination phase*: preparation of protocols and survey material and ethics approval applications.

*Work Package 2: Stakeholder engagement phase, e-surveys and focus groups:* An electronic-survey (e-survey) will be developed by a multi-disciplinary study management group (SMG).

This will be circulated to HCPs involved in the care of neonates and parents/carers whose babies had recently been discharged from hospital. A small number of parents/carers will be invited to take part in focus groups.

*Work Package 3: Co-design of resources and quantitative evaluation*: Parents/carers willing to co-design educational and information resources to support safe administration of medicines to neonates following discharge will be recruited. A quantitative evaluation of the effect of the resource will be conducted with a new group of caregivers to measure specific outcome(s).

**Discussion:** This study aims to co-develop, with healthcare professionals (HCPs) and parents/carers, resources aimed at improving medication safety and reducing parental anxiety for those giving medications to neonates at home. Co-development of resources with HCPs and parents, ensures that the project outcomes are relevant and useful, leading to a reduction in parental anxiety.

**Trial registration:** ISRCTN registry Ref 17332620

## Background

Over 100,000 babies are cared for in neonatal units in the UK and Ireland annually, either because they have been born prematurely, or born full term but require supportive care. This figure represents 1 in 7 babies born in the UK and Ireland each year (1). The majority of preterm infants are admitted onto neonatal units where healthcare staff administer their medicines, however, many neonates remain on medications after they are discharged home requiring parents/carers to administer medicines (1). This is especially true in the case of infants with chronic lung disease, metabolic bone disease, cardiac conditions, congenital hyperinsulinism, endocrine deficiencies, seizures, neonatal abstinence syndrome or neonates undergoing palliative care.

Whilst current practice on most neonatal units in the UK and Ireland is for nursing staff to administer medicines, there has been a recent move towards parents giving medicines in hospital as part of family integrated care (FICare). FICare care has been shown to be beneficial to parents and infants (5) but requires education support from staff. Unfortunately, education about medicines may occur only immediately prior to discharge which can cause high levels of anxiety and concern for parents/carers as they worry about making mistakes including measuring of doses, as they transition to the unsupervised home environment. Added to this anxiety is the fact that premature neonates are often prescribed liquid medications which require small volumes to be measured.

A recent study of babies born prematurely (less than 32 weeks gestation), found that the median number of medicines per patient at discharge was 2.5 (range 2 to 7) with 28% of babies being discharged on more than 3 medicines (1). The frequency that parents and carers administer medicines at home is increasing (6). There are many factors contributing to this trend including; improving survival from serious health problems, the increasing use of medicines to treat health problems, and the desire to move the delivery of care closer to home. A systematic review of carers for all ages estimated the administration error rate at home to be between 2%-33% and that up to 90% of carers administer a medicine incorrectly at some point (7). When parents are responsible for administering medicines to premature infants, an administration error of 66.3% has been reported (7).

Families, clinicians and researchers are aware of the problems related to using medicines in babies and infants at home. A James Lind Alliance priority setting exercise for premature birth has ranked developing packages of support for families at home as the 5th highest out of all topics (3). Further research has shown about a quarter of all families who start a new medicine experience a problem when administering their medicine within the first month (8). There are many sources of difficulty and anxiety faced by parents and caregivers for example, health literacy and language proficiency which has been linked with risk of dosing errors (9). This has been highlighted by UK charity, BLISS, describing difficulties faced by parents/carers of neonates with polypharmacy, unfamiliar devices and dosage regimens (10).

Some of these difficulties could be alleviated through support and education of the caregiver. Parents/carers should be encouraged and supported to participate in their baby’s care on the neonatal unit, and to be involved in discussions and decision-making with the neonatal team. Family Integrated Care (FICare) (5) supports parents’ active involvement in the care of their babies and has a positive impact on both the baby’s and parents’ health and wellbeing (1). This study will be able to support the development of FICare and relieve parental anxiety.

Restrictions due to the COVID pandemic have heightened the need for more extensive support for parents and caregivers during transitions of care. Interactions between HCPs and parents have been reduced, however, the activity on neonatal units has continued. Many units are restricting access to the neonatal unit resulting in access for only one parent who is now the sole decision maker, information gatherer and advocate for their infant. COVID restrictions also mean that parents are not present on the neonatal unit when medication administration is taking place, which can increase pressure and stress on parents.

## Methods/Design

The study design has been planned in accordance with the National Health Service (NHS, UK) NHS and the Health Service Executive (HSE, Ireland), COVID pandemic restrictions reducing physical interactions and using online communication methods where possible. The study is divided into 3 work packages (WP).

### Work package (WP) 1: project management and coordination

Research staff in Alder Hey (AH) Paediatric Medicines Research Unit (PMRU) and Royal College of Surgeons of Ireland (RCSI) will project manage and coordinate the project. Activities include:

- Protocol development and regulatory approvals
- Staff recruitment
- Sponsorship and site(s) set-up
- Budget management
- Ongoing project management (overseen by monthly Study Management Group (SMG) meetings)
- Risk management procedures
- Monitoring of participant recruitment
- Engagement/communication with families through social media platforms with regular updates
- Agree schedule for ongoing report writing and other outputs

The SMG consists of a representative from each study site, the Project Leads and co-applicants in addition to parent representatives and will oversee delivery of tasks in WP1.

### Work Package 2: Stakeholder mapping, development of e-surveys and focus groups

This will generate professional- and parental-informed data on the barriers, challenges and unmet needs of neonatal medicines administration in transitions of care and the home setting. It will include three distinct sub-phases:

1. **Mapping**. Identification of relevant stakeholder groups (e.g. NPPG, BLISS, Irish Neonatal Health Alliance). The first output will be a stakeholder map of institutions, individuals and networks to send the e-surveys.
2. **Development of electronic surveys (e-surveys)**. Customised stakeholder-surveys will be developed using the expertise within the SMG and from the issues highlighted in the background section of this application. E-surveys will contain a combination of open/closed questions, rating scores, etc. Each will be piloted with a sample of the intended respondent groups and adapted if required (to remove ambiguities; estimate data imputation and completeness, etc). Microsoft forms survey software will be used.
  a. **HCPs**. E-surveys will be sent to nurses, doctors, advanced neonatal nurse practitioners (ANNPs) and pharmacy staff in the five study sites. Information regarding the study will be provided via posters at participating sites, via social media and staff networks. The e-surveys will request their views on medication problems encountered by parents/carers during the transition between hospital and home and ask them to identify any available resources used to support this process. E-surveys will also be sent to any additional staff stakeholders and via social media eg Twitter. There is no scientific justification for a target number of respondents to this survey. Based on our previous experience (e.g. Neonatal and Paediatric Pharmacists Group (NPPG) and Modification of Drugs Required in Children (MODRIC) study (12)) we are confident we will obtain a minimum of 75 respondents.
  b. **Parents/Carers**. Parents/Carers groups will be identified via the stakeholder map. Written information about the study will be provided via posters at participating sites, via social media (Instagram, Twitter etc) and parent support groups. The e-survey will request their experience of any problems with medicines or practices they have found beneficial during their transition between hospital and home, again using the issues highlighted by parents including the @LeedsNERDs group as the structure. There is no scientific justification for a target number of respondents to this survey. Based on previous experience (e.g. Survey of Parent administration of medicines on neonatal unit (13)) we are confident we will be able to obtain a minimum of 50 respondents. Paper surveys will be made available upon request - a member of the research team will upload any data from these to the study database.
  The e-surveys will stay open for 4 weeks, from the date that the last study site has opened, with reminders at 1 and 3 weeks after initial contact.
3. **Focus groups**. Outputs from the e-surveys will be used to prepare material for online focus groups with parents/carers to explore the unmet needs identified by parents/carers in more detail. At the end of the e-survey parents/carers will be invited to contact the study team if they are interested in taking part in a focus group. Eligible parent/carers at participating sites will also be invited to take part and given study information to consider. Advertisement for focus group participants will also be shared on social media and via parent support groups. Focus groups will be held at different times on different days in order to increase availability for parents/carers and will be virtual or face to face. Based on previous experience (e.g. Survey of Parent administration of medicines on neonatal unit (13)) we are aiming to recruit a minimum of 25 participants for the focus groups, however, we will stop recruitment to focus groups once data saturation is achieved.

### Work Package 3: Co-design and evaluation of resources

This will be divided into 2 sub-phases:

1. **Construct an educational resource**. Informed by the survey results and focus groups, educational resources will be developed alongside a small group of parents/carers (approximately 10) recruited during WP2. If it is not possible to recruit from parents/carers who have attended the focus groups, we will follow the same recruitment method as previous to recruit new parents willing to help co-design resources. Resources may include items such as a medicine “passport” (which could include background information about the baby’s weight, corrected gestational age, current medication, etc.), tips on how to obtain supply of medicines post-discharge, how to use equipment such as oral syringes, links to parent support groups such as BLISS, etc. The benefit of co-production will include content relevance and specificity, as well as use of language relevant to end-users (parents/carers). The resources will include a co-designed question and answer (Q&A) test for parents/carers which will assess knowledge acquisition for intended users The content can be packaged in a medium which can be flexible and accessible (written/manualised or electronic/digital). For methodological and economic reasons, we will focus on evaluating the content (ensuring no unnecessary expense of digital/App development) therefore it is likely that initially resources developed may be paper-based but our links with the Royal College of Paediatrics and Child Health (RCPCH) MfC board may provide a mechanism to later move to digital options or an App.
2. **Evaluation** We will conduct an evaluation of the resource with a new group of parents/carers (approximately 10-15) to estimate:
  a. <u>Utility</u> - qualitative exploration (short semi-structured interviews) of usability, accessibility, language, detail, size, complexity, etc.
  b. <u>Initial efficacy</u> – Quantitative evaluation of knowledge acquisition. The ‘Q&A test’ will be undertaken with the new group of caregivers before and after use of the resource. This may include, for example, identifying errors in the volumes of medicine drawn up in syringes, identifying which medicine is used for which indication, identifying whether medicines should be taken before or after feeds. It may also include a visual analogue scale to rate their anxiety/confidence in relation to giving medicines at home safely.
  c. <u>Ease of implementation</u> – a questionnaire will be circulated to a small group of HCPs to obtain feedback on training and support required to implement the resources.

This sub-phase represents an early efficacy study of the resources developed using important outcomes identified by both HCPs and parent/carer participants. Identification of these outcomes will be used to inform a larger definitive study if required.

### Study inclusion and exclusion criteria

#### Inclusion Criteria

- Parents/Carers of babies receiving care on a neonatal unit who will require medication to be continued after discharge from hospital
- Parents/Carers whose baby has been discharged from a neonatal unit whose medication has continued after discharge within the last 5 years.

#### Exclusion criteria

- Parents/Carers who do not speak English (the resources will initially be developed in English and then we will apply for further funding to allow them to be translated into other languages)
- Parents/Carers whose baby’s medication is likely to stop prior to discharge
- Parents/Carers whose baby has a terminal diagnosis, a severe congenital abnormality or critical illness (unlikely to survive).
- Parents/Carers less than 16 years of age (the resources will initially be developed for parents/carers able to consent themselves, however, once the co-designed resources are available, we would like to explore utility in this group of parents).

### Sampling

This study aims to co-design resources to improve the safety of neonatal medication administration in the home environment.

We aim to represent the following (if possible) within our total of approx. 25 participants in the focus groups.

- Mothers, fathers and carers
- Range of socio-economic states
- Range of ages
- Single parents/ those in relationships
- Range of ethnicity
- Range of those with first child/subsequent children
- Range of gestational ages of infants e.g. babies who were born preterm vs term.
- Range of duration of NICU stay

We also aim to be able to represent the majority of these groups within the parent/carer groups who co-design and evaluate the resources. We anticipate 300-400 parents/carers per year will be eligible for the proposed study and 25% will agree to take part. Consequently over a 6 month period we expect to recruit 25 patients to focus groups in WP2, 6-8 parents/carers to take part in the co-design of resources and 10-15 will take part in the evaluation of these resources in WP3. Interim monitoring and analyses will be on an ongoing basis.

### Statistical considerations

#### Primary outcome measure

- The level of satisfaction with the co-designed resources elicited by questionnaire and interview by parents/carers
- The proportion of HCPs who think the co-designed resources are useful and easy to implement

#### Secondary outcome measures

- Development of a stakeholder map of institutions, individuals and networks
- Produce a list of issues based on outputs from electronic surveys and parent/carer focus-groups and develop into themes
- Produce a list of available resources to support parents/carer administer medicines to neonates and evaluate their contents/structure
- Produce co-designed educational resources based on feedback and parent/carer co-design
- Qualitative assessment of the usability and accessibility of co-designed resources by a new parents/carers
- Quantitative assessment of efficacy of the co-designed resources by a new group of care-givers
- Assessment of the ease of implementation by HCPs

### Analysis Plan

As this is exploratory work no formal hypothesis testing will be undertaken.

#### Analytical approach - Qualitative components

For the qualitative data we will use both an inductive and interpretive approach, and analysis will be informed by the Framework approach (14,15), intended to develop a thematic analysis of spoken data derived from focus groups. Framework Analysis (FA) was developed by Ritchie and Spencer for policy research but is now used widely in other areas including health research (16). There are 5 main stages of FA: familiarisation through immersion in the data; developing a theoretical framework by identifying recurrent and important themes; indexing/coding and charting; summarising data in analytical framework and synthesising data by mapping and interpreting. Its systematic approach makes it suitable for involving all members of a multi-disciplinary team. An open, critical and reflexive approach from all team members is essential

#### Quantitative analysis

Quantitative data will be reported using descriptive statistics, mean/medians for continuous data and counts/percentages for categorical data. To assess variability and uncertainty all outcome measures will be presented with 95% confidence intervals. A more detailed statistical analysis plan will be written when the exact nature of the data is known. Data analysis will be ongoing throughout the study.

### Ethics

Ethical approval has been sought and granted from an approved NHS ethics committee NRES London - Bloomsbury Research Ethics Committee and HRA and Health and Care Research Wales (HCRW) (reference: 21/LO/0351). Ethical approval was sought and granted from the ethics committee at the Rotunda Hospital, Dublin, Ireland : REC-2021-O13.The study will be conducted according to the principles of UK Policy Framework for Health and Social Care Research (17).

### Dissemination plan

The main output from this study will be co-produced parent/carer information that is standardised and in an appropriate format that will allow it to be used across all neonatal units in the UK and Ireland. We will collaborate with Medicines for Children (MfC), and plan to submit this material to them for consideration to publish on their website as a mechanism for dissemination.This will also support the review and updating of the information. BLISS also provides training for neonatal staff around discharge support and the information will be shared with them.

We will publish findings from this study in peer reviewed publication(s) and will produce a written report for the funder as directed by their terms and conditions of the award as funder of the project.

In addition to peer reviewed publications, posters and/or oral presentations will be submitted to leading national conferences of stakeholder groups. This will include the British Association of Perinatal Medicines, Royal College of Paediatrics and Child Health, and the Neonatal and Paediatric Pharmacists Group. Where possible, presentations will be video recorded and uploaded to YouTube. Families will be invited to participate in these activities.

To supplement this research output, we will write commentary articles in the form of blog posts, to coincide with the publication of results. These will be for publication on the NPPG, Pharmaceutical Journal (as non-peer reviewed opinion pieces), and the Leeds Hospitals/AH/Liverpool Women’s/Rotunda/Wirral Research and Innovation websites.

Project updates will be posted in electronic form to social media accounts (e.g. Facebook, Instagram, Twitter) to publicise the research more widely to the public.

## Discussion

PADDINGToN is the first large multicenter study evaluating parent and carer experience in the administration of medicines following discharge of neonates. Several studies have reported high levels of anxiety and concern for parents/carers as they worry about making mistakes including measuring of doses, as they transition to the unsupervised home environment. The aim of the study is to identify through consultation with HCPS and parents and carers the main challenges faced by parents and carers following discharge of their baby from hospital. Using this information the researchers will co-develop, with healthcare professionals (HCPs) and parents/carers, resources aimed at improving medication safety and reducing parental anxiety for those giving medications to neonates at home.

From inception, PADDINGToN has had strong involvement and support from HCPs, parents, carers, the Medicine for Children (MfC) project and parent support groups. The project protocol was written in conjunction with parent representatives who are members of the project management group. It is believed that through extensive PPI the resources developed will be relevant and useful leading to a decrease in parental anxiety.

## Data Availability

No datasets were generated or analysed during the current study. All relevant deidentified data from this study will be made available upon study completion.

## List of abbreviations

AH: Alder Hey
ANNP: advanced neonatal nurse practitioner
FA: Framework Analysis
FICare: family integrated care Healthcare professionals
(HCPs) HSE: Health Service Executive
MfC: Medicines for Children
MODRIC: Modification of Drugs Required in Children
NHS: National Health Service
NPPG: Neonatal and Paediatric Pharmacists Group
PMRU: Paediatric Medicines Research Unit
RCSI: Royal College of Surgeons in Ireland
SMG: study management group
WP: work packages

## Declarations

### Ethics approval and consent to participate

Electronic study data will be kept on encrypted, secure computers or devices within the Lead site. Electronic documents will be used as much as possible, however, if paper documents are used (e.g. consent forms), the originals will be scanned to an electronic site file (eISF) which will be stored within the Trusts secure network. The original paper copies will then be destroyed. The confidentiality of all study participants will be maintained in accordance with the EU GDPR and the Data Protection Act 2018 and data will be held and handled according to Caldicott principles.

Written informed consent from focus group participants will be obtained and recorded using the appropriate study documents. Participants will be free to withdraw from the study at any time and do not need to provide a reason for doing so. Any data collected from participants prior to withdrawal, will be destroyed upon request.

Data and all study documentation will be stored for a minimum of 10 years after the completion of the study, including the follow-up period, unless otherwise directed by the sponsor/regulatory bodies. Once this study has ended and is ready to be archived, the eISF will be stored on the Trust secure network. This may include some personal identifiable data (such as participants’ full name). Participants will be made aware of this process.

## Consent for publication

Not applicable

## Availability of data and materials

The datasets used and/or analysed during the current study are available from the corresponding author on reasonable request

## Competing interests

The authors declare that they have no competing interests

## Funding

This work was financially supported by a Neonatal and Paediatric Pharmacists Group (NPPG) Grant 2020. The funders have had no part in the in the design of the study and collection, analysis, and interpretation of data and in writing the manuscript.

## Authors’ contributions

Project was conceived, designed and developed by: AG, LB and FOB The protocol was written by: AG, LB and FOB

Expert support during the development and writing of the protocol was given by: CB, NC, BC, NM, SM, EN, MT, MP.

All authors read and approved the final manuscript

## Acknowledgements

We would like to acknowledge Rachel Corry and Julie Murphy as parent representatives on our Study Management Team for their valuable contribution to the PADDINGToN project.

## Notes

### Competing Interest Statement

Andrea Gill and Neil Caldwell both have non-financial competing interests as committee members of the funding body NPPG. However, they were excluded from the grant application and review process, which was peer reviewed. The rest of the authors have declared that no competing interests exist.

### Funding Statement

This study is funded by the Neonatal and Paediatric Pharmacists Group UK. https://nppg.org.uk/ LB, AG and FOB were the recipients of the grant. The funders had and will not have a role in study design, data collection and analysis, decision to publish, or preparation of the manuscript.

### Author Declarations

Ethical approval has been sought and granted from an approved NHS ethics committee NRES London - Bloomsbury Research Ethics Committee and HRA and Health and Care Research Wales (HCRW) (reference: 21/LO/0351). Ethical approval was sought and granted from the ethics committee at the Rotunda Hospital, Dublin, Ireland : REC-2021-O13

